# Efficacy and Safety of Metronomic Capecitabine in Hepatocellular Carcinoma: A Systematic Review and Meta-Analysis

**DOI:** 10.1101/2024.06.08.24308644

**Authors:** Nandini Gupta, Neelkant Verma, Bhoomika Patel

## Abstract

**Background and Objective:** Metronomic capecitabine has been found to be useful in several types of cancers such as pancreatic cancer, breast cancer, gastrointestinal cancers, nasopharyngeal carcinoma, metastatic colorectal cancer etc. This unique systematic literature review and meta-analysis was undertaken to assess the effectiveness and safety of metronomic capecitabine as a treatment regimen for hepatocellular carcinoma.

**Method:** A systematic search of major databases was performed. Eight studies that dealt with the use of metronomic capecitabine for HCC were selected, seven were non-RCTs, and one was an RCT. Meta-analysis of these studies was performed using Review manager v5.3 and STATA 15.1 software. The pooled prevalence of OS, PFS, ORR, Grade 1-2 AE, Grade 3-4 AEs were determined, including publication bias and sensitivity analysis.

**Result:** Eight studies met the inclusion criteria, combining the pooled data of 476 patients from safety and efficacy studies. The pooled prevalence of DCR and ORR achieved with metronomic capecitabine were 36% (95% CI 32-41) and 7% (95% CI 5-9) respectively. The median PFS and median OS were 3.57 months (95% CI 3.29-3.85) and 11.75 months (95% CI 10.56-12.95) respectively. The incidence of 3-4 grade AEs and 1-2 grade AEs were 38% (95% CI 32-44) and 73% (95% CI 67-79) respectively.

**Conclusion:** This meta-analysis highlights metronomic capecitabine as a potential second-line therapy for HCC. However, effective management of capecitabine’s side effects is essential.

## **1.** Introduction

Hepatocellular carcinoma (HCC) is the deadliest form of liver cancer, typically arising from chronic liver disease and cirrhosis. It is the third most frequent cancer encountered worldwide ^1^. According to the most recent data from GLOBOCON 2020, liver cancer is the third leading cause of cancer-related deaths in 46 nations, exhibiting notably high fatality rates. In 2020, 905,700 cases of liver cancer were reported worldwide, resulting in 830,200 fatalities ^2^. According to reports by the World Health Organization, by 2040, the number of new cases and deaths from liver cancer is expected to increase by >55% ^2^. The five-year survival rate of all SEER (Surveillance, Epidemiology, and End Results) stages combined was found to be 21% ^3^.

Current treatment options available for treating hepatocellular carcinoma depends on which stage the hepatocellular cancer is diagnosed. At present, several treatment options are available for early detection, such as liver transplantation, surgical resection, and ablation therapy. If HCC is detected at an intermediate stage, radiation therapy, chemotherapy, and embolization therapy are available. During the advanced stages of hepatocellular cancer, targeted therapies, such as molecular targeted therapy, immunotherapy, and gene therapy, are administered to patients ^4^.

Chemotherapy is considered to be a mainstay treatment available for advanced HCC. The first- and second-line chemotherapeutic agents used are sorafenib, sunitinib, brivanib, linifanib, erlotinib, doxorubicin, ramucirumab, tivantinib etc.^5^. Traditional agents of chemotherapy used viz. cyclophosphamide, oxaliplatin, gemcitabine, cisplatin, methotrexate etc. were found to be associated with certain patterns of hepatotoxicity. Traditional chemotherapeutic agents, such as cyclophosphamide, oxaliplatin, gemcitabine, cisplatin, and methotrexate, are associated with certain patterns of hepatotoxicity. These toxicity patterns are characterized by sinusoidal obstructive syndrome, pseudocirrhosis, steatosis, acute hepatitis, and hepatic necrosis ^6^. Chemotherapy also results in resistance to signaling pathway activation, hypoxia, and genetic abnormalities, further deteriorating the chemotherapy regimen ^7^.

Unlike traditional chemotherapy, metronomic therapy (MCT) utilizes low, frequent doses of medication delivered continuously or regularly (daily or weekly) over an extended period, aiming to minimize side effects ^8^. These low doses are usually 1/10^th^-1/3^rd^ of the maximum tolerated dose of certain anti-neoplastic drugs which thereby enhances the anti-angiogenic property of the drugs ^9^. When given at small dosages and frequently, metronomic medications have the potential to produce a prolonged low blood level of the medication without causing noticeably harmful side effects ^10^. Metronomic chemotherapy presents a compelling alternative due to its triple benefit: improved tolerability (better toxicity profile), reduced financial burden (lower cost), and simplified treatment approach (easier use and administration) compared to conventional regimens ^11,12^.

Capecitabine (CAP) is a prodrug of 5-florouracil chemotherapeutic drug. It causes cell cycle arrest and apoptosis by blocking DNA polymerase ^13^. This drug is usually administered orally ^14^. Various clinical trials have reported that use of MCT, MCT combined with other treatments (such as targeted therapies, biologics, and endocrine therapy) has chances of better overall survival of the patients ^15^.

Several studies have been performed involving the use of metronomic capecitabine on metastatic breast cancer ^16^, pancreatic cancer ^17^, and various other cancers. The integration of metronomic capecitabine with this drug regimen has demonstrated enhanced overall survival and effectiveness among patients. This meta-analysis and systematic literature review highlights how metronomic capecitabine can improve results for individuals diagnosed with hepatocellular carcinoma.

## **2.** Method

To ensure data clarity and prevent any potential ambiguities, we registered the study protocol in the International Prospective Register of Systematic Reviews (PROSPERO, CRD42023415892). Preferred Reporting Items for Systematic Review and Meta-analysis (PRISMA) guidelines were followed to conduct the current meta-analysis.

### 2.1. Extraction of data from various sources

Various databases such as PubMed, Scopus, Cochrane library, ProQuest and Science Direct were independently searched by two investigators to look for the published studies till 21^st^ March 2023. The search strategy used the fixed keywords search with “metronomic capecitabine” AND “cancer”.

### 2.2. Inclusion criteria

The study included patients who met the following criteria (1) Retrospective and prospective studies including randomized and non-randomized controlled studies using metronomic capecitabine for treatment of hepatocellular cancer of any stage. The studies that are in English language were selected. (2) The study examined various outcomes, including progression-free survival (PFS), disease control rate (DCR), overall response rate (ORR) and overall survival (OS), either as reported or derivable from the data at hand. (3) Research papers which included the presence of chemotherapy-induced severe adverse events evaluated at grade 1/2 or higher.

### 2.3. Exclusion criteria

The study excluded patients who met the following criteria (1) Studies where capecitabine chemotherapy was given concurrently with other treatment approaches like immunotherapy, radiotherapy, or targeted therapy. (2) Fundamental research (non-clinical studies), redundant publications, individual case reports, review papers and studies from which pertinent data couldn’t be retrieved were excluded.

### 2.4. Data extraction

The search method was determined from the combined efforts of all authors. Two investigators independently selected eligible studies, searched the literature, and extracted data. Mutual discussions were conducted if disparity arose at any point during the study. We collected information on the study’s characteristics, study phase, study location, study duration, number of patients, gender distribution, median patient age, ECOG score, diagnostic criteria, inclusion-exclusion criteria, prior chemotherapy treatments, 3-4 grade AEs, 1-2 grade AEs, overall response rate (ORR), disease control rate (DCR), progression-free survival (PFS), and overall survival (OS) (Table 1).

**Table 1:**
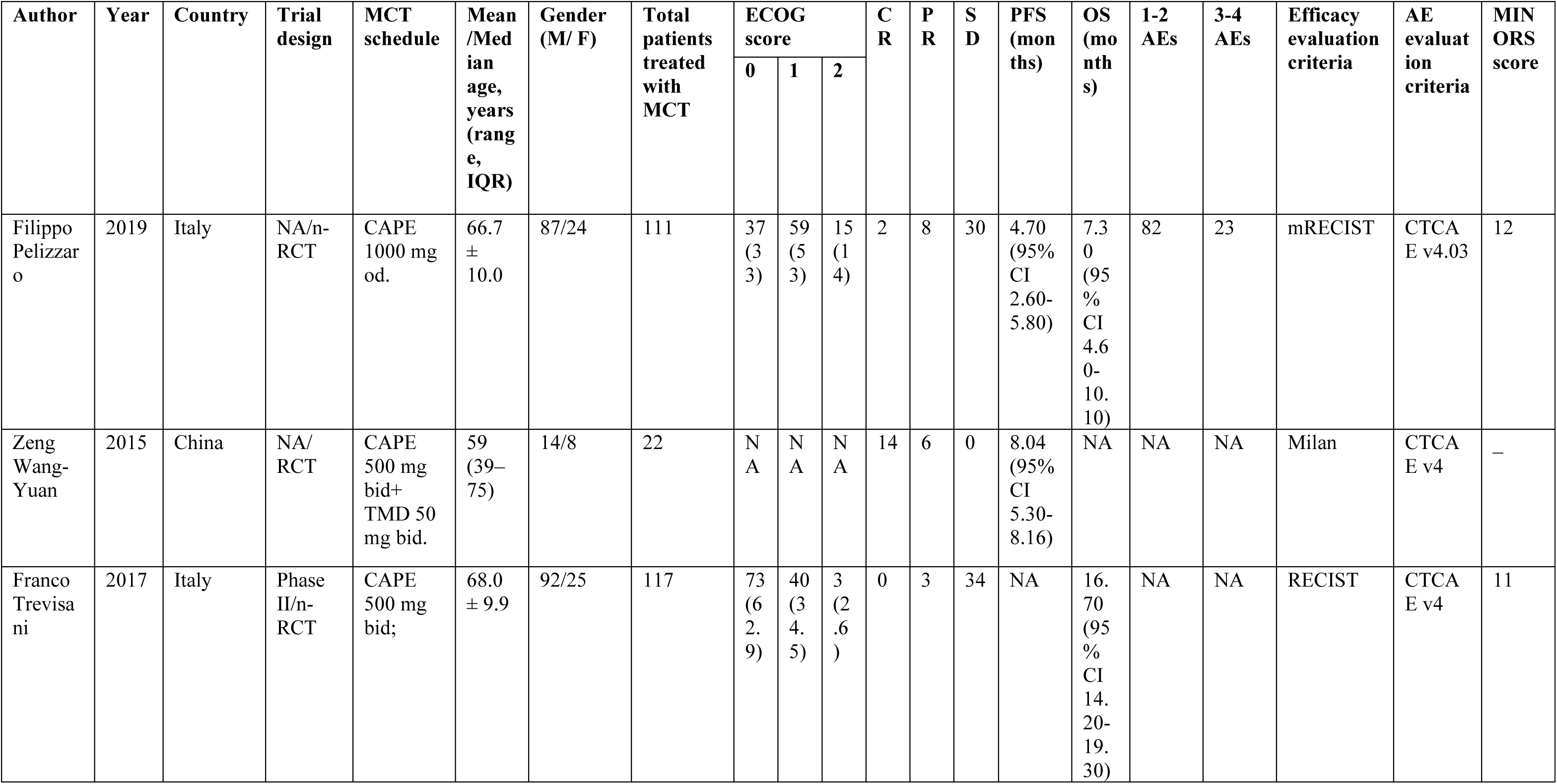
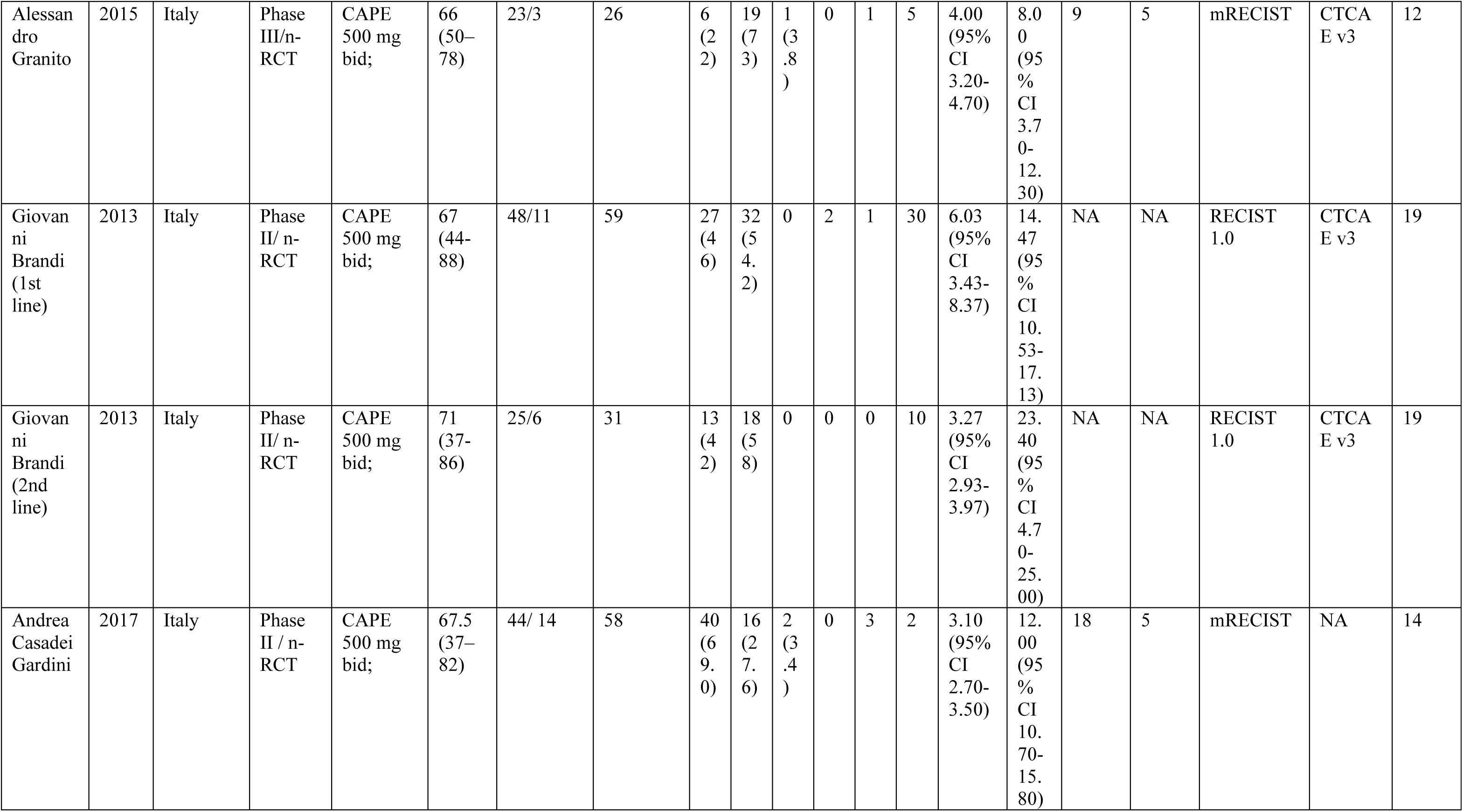
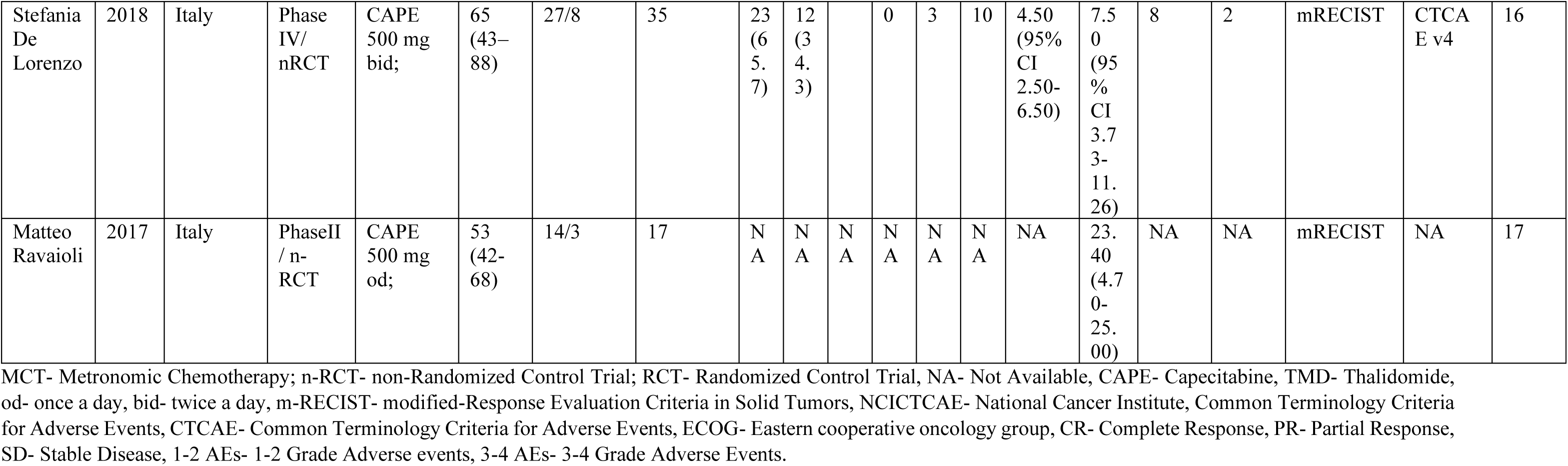
Basic information and definite characteristics of the eligible studies.

### 2.5. Quality evaluation

Quality assessment of the eight included studies was independently performed by two researchers. The evaluation utilized the Methodological Index for Non-randomized Studies (MINORS) tool for non-RCTs, and the Cochrane risk of bias tool for the studies was used for randomized controlled trials (RCTs).

### 2.6. Statistical evaluation

A metronomic capecitabine regimen used for hepatocellular carcinoma as second-line treatment is presented through this meta-analysis and the pooled prevalence is reported. A range of study types, encompassing randomized controlled trials (RCTs), single-arm studies, and non-RCTs, were amalgamated to calculate the collective occurrence rates of DCR, ORR, PFS, OS, 3-4 grade AEs, 1-2 grade AEs. For studies lacking reported data, we used Webplotdigitizer version 3.8 to extract the data from relevant graphs. The analysis combined data using methods specific to the type of outcome. For odds ratios (OR) and proportions associated with ORR, DCR, and AE incidence, a ratio merging method incorporating confidence intervals (CIs) was employed. Conversely, median PFS and OS were estimated using a mean merging method that considered both upper and lower bounds of the data and their respective CIs. To assess variability (heterogeneity) between studies within each outcome group, I² statistic and Cochran’s Q were utilized. When significant heterogeneity was detected (p-value < 0.10 or I² > 50%), a random-effects model was implemented for meta-analysis. This model accounts for potential variations in treatment effects across studies. The influence of any single study on the combined outcome was evaluated through sensitivity analysis, where the overall results were re-evaluated by sequentially excluding each study one at a time. Finally, potential publication bias, where unpublished studies with negative or non-significant findings might be missing, was assessed using funnel plots. A p-value less than 0.05 was considered indicative of publication bias. All statistical analyses were performed using STATA 15.1 software (Stata Corp LP, College Station, TX, USA).

## 3. Results

### 3.1. Number of studies

A total of 305 number of articles were retrieved through the database search. This systematic search retrieved 21 articles from PubMed, 60 from Scopus, 32 from Cochrane library, 148 from ProQuest and 44 from Science Direct. After the removal of 34 duplicates from the Mendeley reference manager software, a free citation tool, the total number of articles left for screening were found to be 271. A total of 221 articles were removed from the screening as they were not able to meet the inclusion and exclusion criteria which is explained in Figure 1. Therefore, a total of 50 studies were assessed and met the eligibility criteria. However, following a comprehensive review of these 50 articles, 42 of them were deemed ineligible for the study. This was primarily due to variations in their study protocols, the involvement of drugs other than capecitabine, and the presence of some review-type articles. Out of the selected eight studies, one is RCT i.e. study conducted by Zeng et. al., ^18^ and others are non-randomized clinical trial, Alessandro et. al.^19^, Stefania et. al.^20^, Andrea et. al. ^21^, Giovanni et. al. ^22^, Filippo et. al. ^23^, Franco et. al. ^24^ and Matteo et. al., ^25^. These all 7 non-RCT studies were considered for pooling data so that further analysis can be performed. PRISMA Flowchart for the retrieval of article and selection is given by Figure 1.

**Figure 1:**
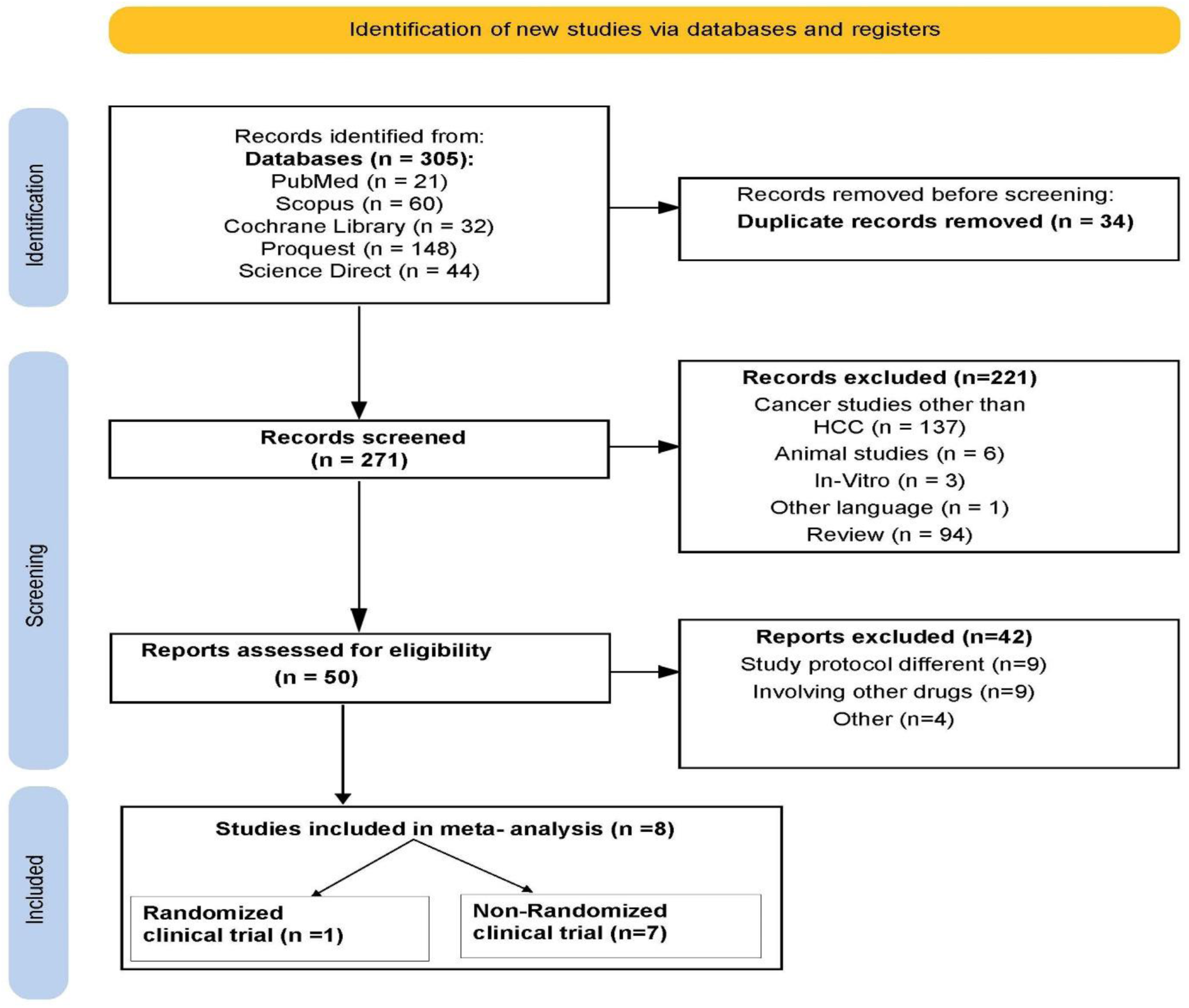
Step-by-step followed in the study from article retrieval and selection.

### 3.2. Quality of studies

The evaluation of scores in RCT showed a good value in Cochran’s risk of bias assessment, but, some unclear risk of selection bias was found in the study done by Zeng et al.^18^ . Figure 2 (a) indicates risk of bias graph and (b) indicates risk of bias summary.

**Figure 2:**
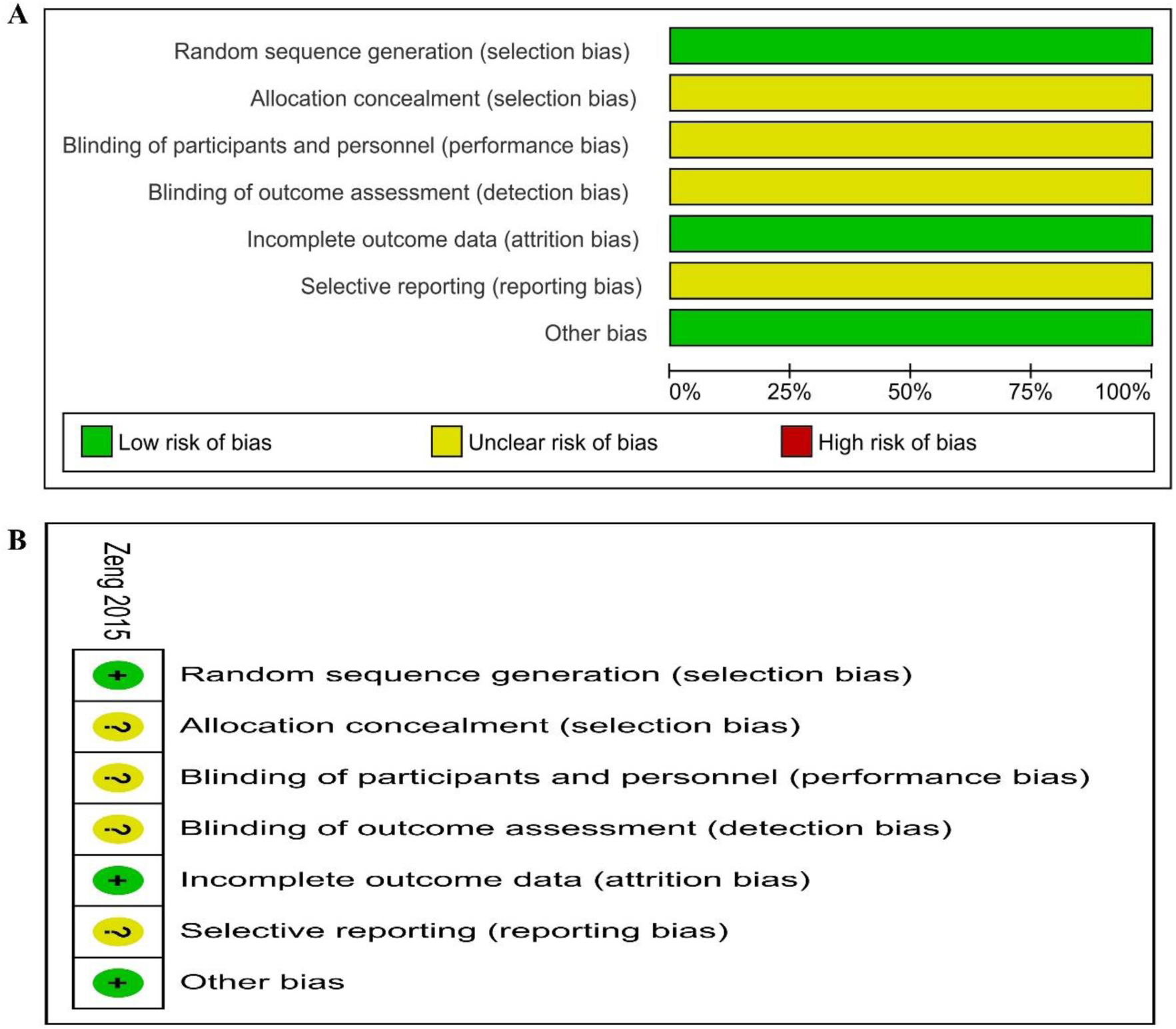
Assessment of risk of bias for included Randomized Clinical Trials (RCTs) to determine the quality of included RCTs using the Cochrane risk of bias method. (a) Risk of bias graph: review authors’ judgments on each risk of bias item presented as percentages across all included studies. (b) Risk of bias summary: review authors’ judgments on each risk of bias item for each included study.

All the single-arm studies scored good scores (between 11-19) which suggests good quality studies (Table 1). The mean age of the patients was reported in two studies and they were found to be 66.7 ± 10.0 in the study done by Filippo et. al. ^23^ and in the study by Franco et. al. ^24^, the mean age is reported as 68.0 ± 9.9 years respectively. In the rest of the studies, the median age was reported and they all were found to be between 53-68 years. Mostly in all the studies, ECOG score was reported and was found to be between 0-2 in the studies by Alessandro et. al.^19^, Stefania et. al.^20^, Andrea et. al. ^21^, Giovanni et. al. (1^st^ and second line), ^22^, Filippo et. al. ^23^, and Franco et. al. ^24^. ECOG scores were not reported in two studies ^18,25^.

### 3.3. Combined data analysis

We analyzed data from eight studies involving a total of 476 patients. Eight of these were clinical trials, some looking back at existing data (retrospective) and others following patients over time (prospective). There was one additional study by Giovanni et. al. ^22^ that investigated both first-line and second-line treatments for the same condition. Since this study analyzed these treatments separately, we treated it as two studies for our analysis. Detailed PFS were found to be in seven studies and the detailed PFS was extracted from Zeng et. al. ^18^ using Webplotdigitizer version 3.8 tool from the graph provided in the paper. The detailed OS data was available in eight studies, the median PFS and OS were 3.57 months (95% CI 3.29-3.85) and 11.75 months (95% CI 10.56-12.95) respectively.

Four studies reported incidence of 1-2 grade AEs and 3-4 grade AEs (Table 1).

The incidence of grade 3-4 AEs and grade 1-2 AEs were 38% (95% CI 32-44) and 73% (95% CI 67-79) respectively. The 3-4 grade AEs included fatigue, 5% (95% CI 2-7), bilirubin elevation/ liver toxicity, 5% (95% CI 1-10), anemia, 4% (95% CI 2-6), hand foot reaction, 2% (95% CI 0-3) and thrombocytopenia, 2% (95% CI 1-4). Grade 1-2 AEs included fatigue, 56% (95% CI 51-60), thrombocytopenia, 10% (95% CI 7-13), hand foot reaction, 7% (95% CI 5-9), anemia, 7% (95% CI 5-9), bilirubin elevation/ liver toxicity, 7% (95% CI 3-11), nausea/ vomiting, 5% (95% CI 2-7) and diarrhea, 3% (95% CI 1-5) (Figure 3, Table 2 and Table 3).

**Figure 3:**
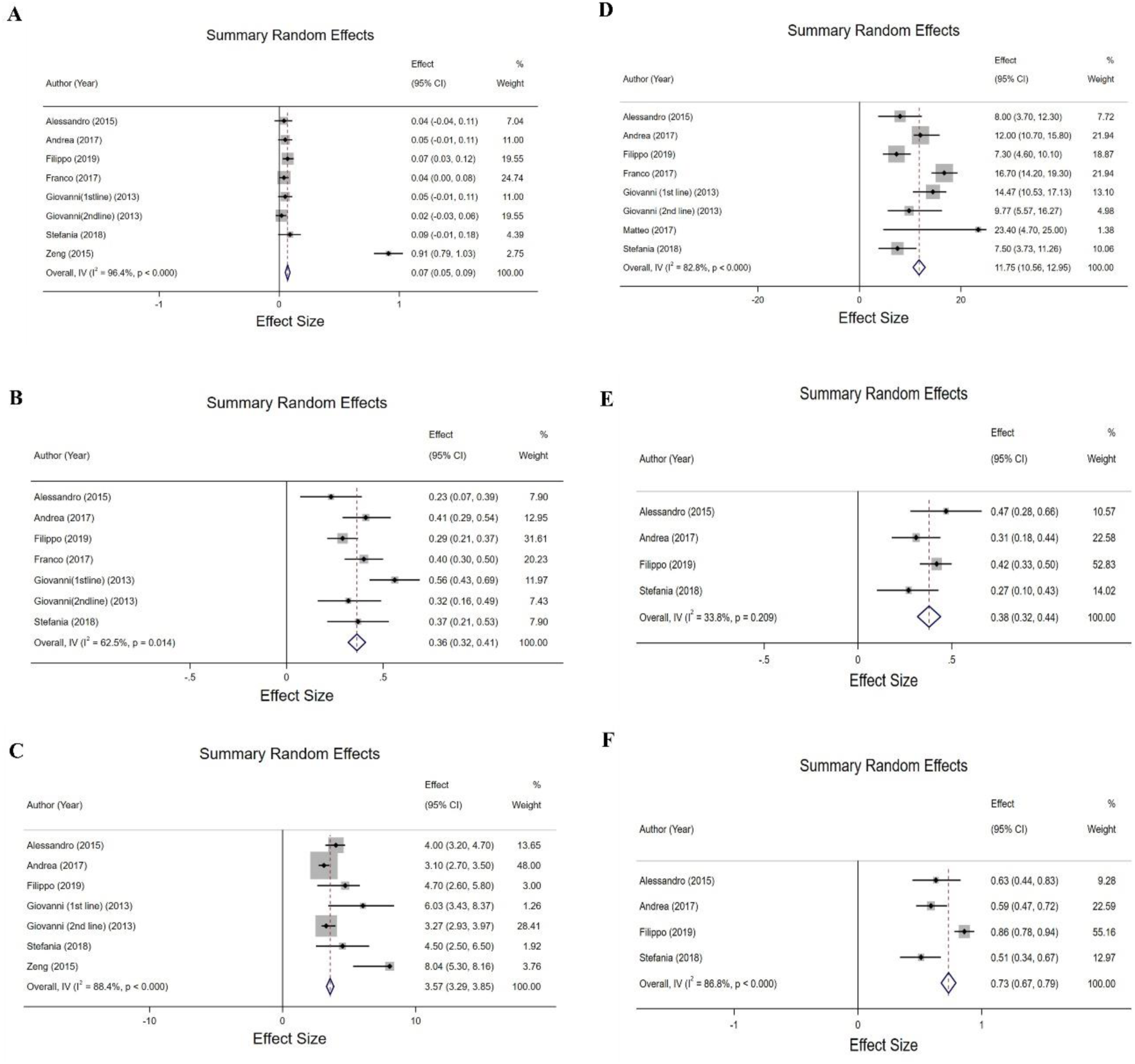
ORR, DCR, PFS, OS, Grade 3-4 AE, Grade 1-2 AE incidence of metronomic capecitabine (a) ORR (b) DCR (c) PFS (d) OS (e) Grade 3-4 AE (f) Grade 1-2 AE.

**Table 2:** Incidence of specific and grade 1-2 AEs caused by metronomic oral capecitabine monotherapy.

|  | **No. of**<br>**clinical**<br>**trials** | **Incidence%**<br>**(95% CI)** | **I<sup>2</sup> %** | **p value** |
| --- | --- | --- | --- | --- |
| **Anemia** | 7 | 7 (5-9) | 86.5 | <0.000 |
| **Hand Foot Reaction** | 9 | 7 (5-9) | 52.4 | 0.032 |
| **Thrombocytopenia** | 6 | 10 (7-13) | 89.3 | <0.000 |
| **Fatigue** | 7 | 56 (51-60) | 48.8 | 0.069 |
| **Nausea/ Vomiting** | 4 | 5 (2-7) | 23.5 | 0.27 |
| **Diarrhea** | 4 | 3 (1-5) | 33.9 | 0.209 |
| **Bilirubin elevation/ liver**<br>**toxicity** | 5 | 7 (3-11) | 66.8 | 0.017 |

**Table 3:** Incidence of specific and grade 3-4 AEs caused by metronomic oral capecitabine monotherapy.

|  | **No. of**<br>**clinical**<br>**trials** | **Incidence%**<br>**(95% CI)** | **I<sup>2</sup> %** | **p value** |
| --- | --- | --- | --- | --- |
| **Anemia** | 5 | 4 (2-6) | 0 | 0.574 |
| **Hand Foot Reaction** | 4 | 2 (0-3) | 0 | 0.715 |
| **Thrombocytopenia** | 5 | 2 (1-4) | 0 | 0.453 |
| **Fatigue** | 3 | 5 (2-7) | 16.9 | 0.3 |
| **Bilirubin elevation/ liver**<br>**toxicity** | 3 | 5 (1-10) | 0 | 0.519 |

Eight studies evaluated the objective response rate (ORR), which refers to the percentage of patients who achieved complete or partial tumor shrinkage. Seven studies assessed the disease control rate (DCR), encompassing both complete/partial responses and stable disease. The DCR and ORR achieved with metronomic capecitabine were 36% (95% CI 32-41) and 7% (95% CI 5-9) respectively.

### 3.4. Assessment of publication bias and sensitivity analysis

A visual evaluation of the funnel plot and the results of Egger’s test were combined to examine the possible impact of publication bias on our meta-analysis findings. The results of Egger’s test (ORR, DCR, PFS, OS, Grade 3-4 AEs, Grade 1-2 AEs) did not show any publication bias (p=0.070, p=0.708, p=0.041, p=0.991, p=0.639, p=0.140). The funnel plots also did not show any remarkable asymmetry, suggesting that publication bias may not be a major concern in our analysis (Figure 4).

**Figure 4:**
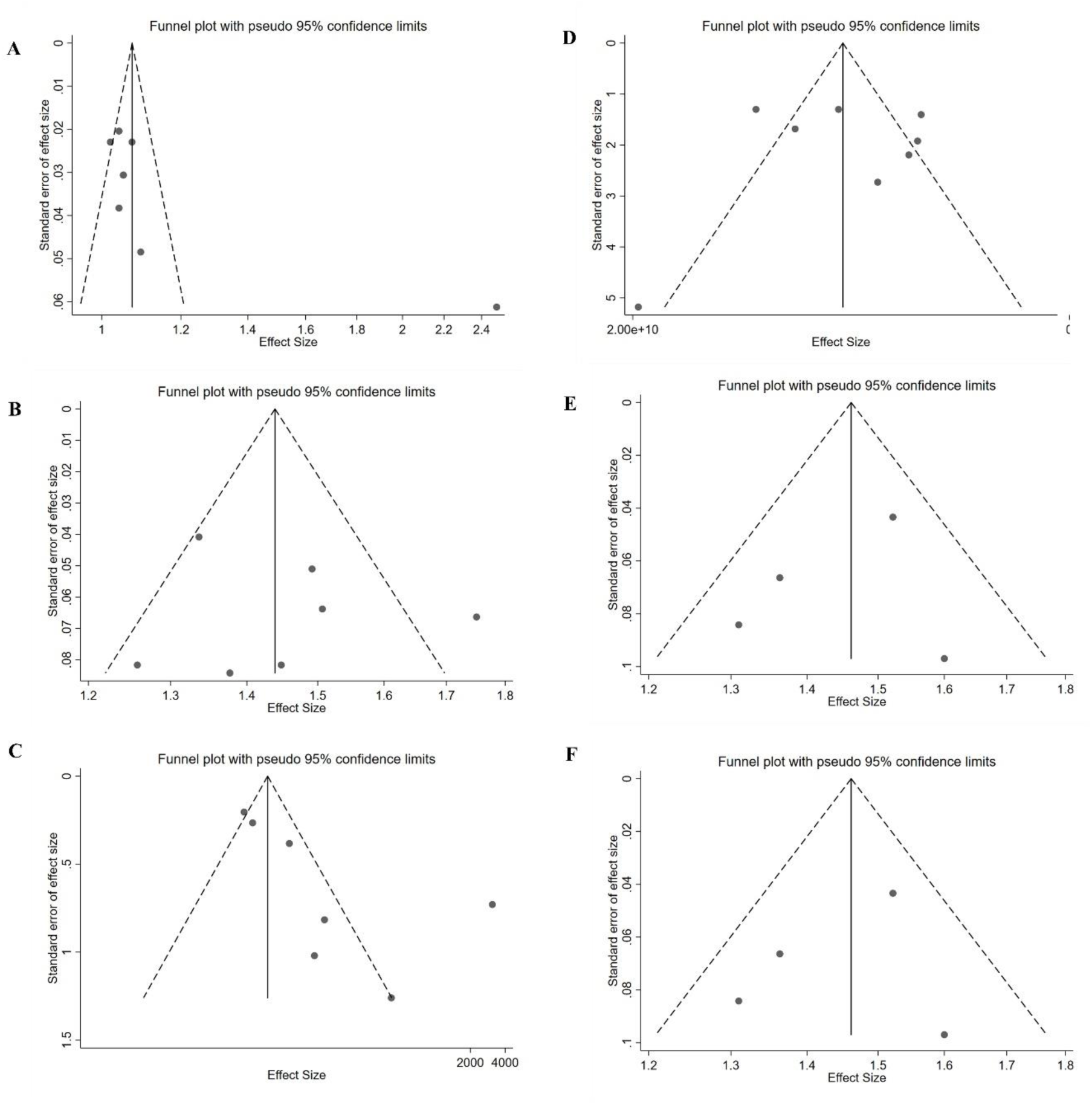
Funnel plot for assessing publication bias of ORR, DCR, PFS, OS, Grade 3-4 AE, Grade 1-2 AE incidence of metronomic capecitabine (a) ORR (b) DCR (c) PFS (d) OS (e) Grade 3-4 AE (f) Grade 1-2 AE.

To assess the influence of any single study on the combined outcome, we conducted a sensitivity analysis where the overall results were re-evaluated by sequentially excluding each study one at a time. The overall effect size remained statistically significant and directionally consistent with the original analysis (Figure 5).

**Figure 5:**
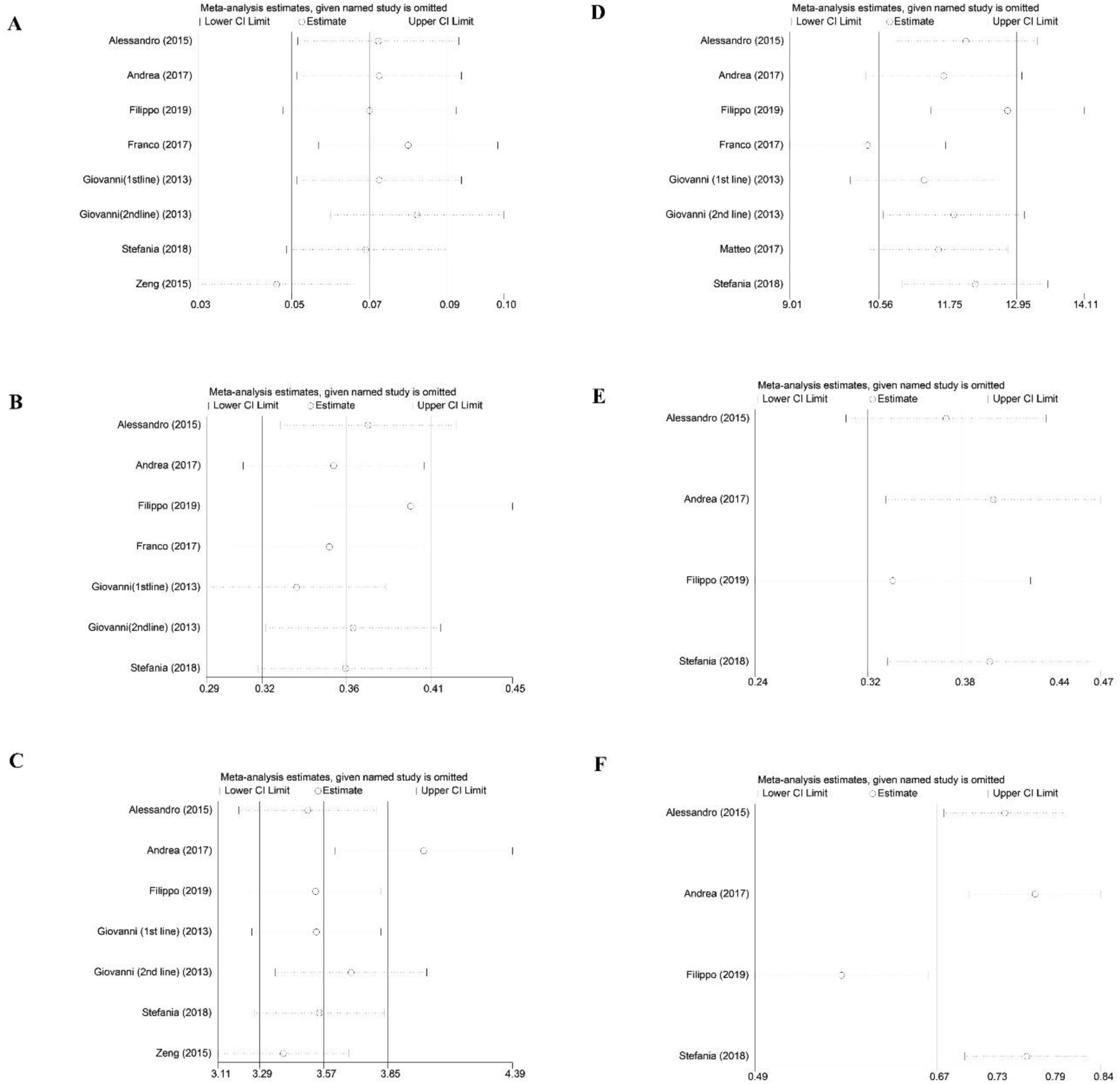
Sensitivity analysis tested the effects on overall results omitting one study at a time of ORR, DCR, PFS, OS, Grade 3-4 AE, Grade 1-2 AE incidence of metronomic capecitabine (a) ORR (b) DCR (c) PFS (d) OS (e) Grade 3-4 AE (f) Grade 1-2 AE.

## 4. Discussions

Hepatocellular carcinoma is characterized through the complex multistep process. In the recent years, significant progress has been made in the treatment of hepatocellular carcinoma, especially in the field of targeted treatments and immunotherapies. The continuous administration of low-dose chemotherapy drugs administered over an extended period of time is known as metronomic chemotherapy (MCT) ^26^. Metronomic chemotherapy is administered in various types of cancer malignancies such as in the gastrointestinal tract, respiratory system, blood, brain, skin, and genitourinary systems, breast and various other cancers ^27^. The main benefits of metronomic chemotherapy are better toxicity profile, lower cost, easier use and administration over conventional chemotherapeutic regimens ^11,12^. The metronomic chemotherapeutic drugs impair the cells responsible for the angiogenic mechanisms thereby rendering the tumoral endothelial cells in overcoming the drug resistance ^12^.

Numerous research studies have explored the use of metronomic chemotherapy in the context of hepatocellular carcinoma. A meta-analysis was conducted on the patients suffering from stage-C hepatocellular carcinoma with poor liver function. The two-drug regimen comparison was carried out on sorafenib and a combination of metronomic drugs inclusive of epirubicin, cisplatin and 5-flourouracil. It was concluded from the study that MCT is a good alternative treatment option especially in patients with poor liver function^28^. A complete remission of a diffuse large B-cell lymphoma was observed in a young patient when treated with metronomic chemotherapy involving combination of drugs of prednisone, vinorelbine, etoposide, cyclophosphamide, plus rituximab and finally ibrutinib^29^.

Capecitabine which is a fluoropyrimidine carbamate drug which is used since more than 20 years for the treatment of various cancers such as head and neck cancers, gastrointestinal cancers, urinary tract cancers and breast cancer, colorectal cancer etc. ^30^. Capecitabine is administered orally and is rapidly absorbed and taken up from the gut ^31^. The specific dosage and treatment regimen for capecitabine depend on the type and stage of cancer being treated, as well as the patient’s individual health and tolerance to the medication. It is typically taken in cycles, with periods of treatment followed by rest periods ^32^. Through this study, we aim to bring insights for the clinicians for selection of metronomic capecitabine for the treatment of HCC.

Quality evaluation was performed in our meta-analysis using the Cochran’s risk of bias assessment and it was found that the evaluation of scores in RCT showed a good value but, some unclear risk of selection bias was found in one of the studies ^18^. All the single-arm studies showed scored good scores (between 11-19).

In current study, median OS and PFS were 11.75 months (95% CI 10.56-12.95) and 3.57 months (95% CI 3.29-3.85) respectively. The DCR and ORR achieved were 36% (95% CI 32-41) and 7% (95% CI 5-9) respectively. A study carried out in metastatic colorectal cancer patients involved use of metronomic capecitabine as maintenance treatment. The median overall survival and median progression-free survival were observed to be 23.82 (95% CI 22.38–25.25) months and 5.66 (95% CI 5.25–6.07) months respectively ^33^. In another study, conducted on patients with metastatic neuroendocrine tumors involving drug regimen of bevacizumab, octreotide and metronomic capecitabine, the median PFS was 14.9 months ^34^. Likewise, a retrospective analysis of metronomic capecitabine in metastatic gastro entero-pancreatic neuroendocrine tumors was conducted. The progression-free survival was found to be 9.9 months ^35^. Metronomic capecitabine was administered in metastatic breast cancer patients and its efficacy and safety were assessed. Median OS was found to be 17 months ^36^. In another study, patients in the SYSUCC-001 trial suffering from early-stages of triple-negative breast cancer were evaluated to check the good effect of metronomic capecitabine. The predicted pooled analysis of 5-year OS was 85% ^37^. Likewise, a meta-analysis conducted by Liu et al. examined the effectiveness of various metronomic chemotherapy regimens involving capecitabine, methotrexate, vinorelbine, bevacizumab etc. and its combinations for metastatic breast cancer. The 12-month overall survival (OS) and 6-month progression-free survival (PFS) rates were determined to be 70.3% (95% CI 62.6–76.9) and 56.8% (95% CI 48.3–64.9) respectively^38^.

To assess the safety and toxicity of metronomic capecitabine in hepatocellular carcinoma, a meta-analysis was performed. The incidence of grade 3-4 AEs and grade 1-2 AEs reported in our study were 38% (95% CI 32-44) and 73% (95% CI 67-79) respectively. Incidence of Grade 1-2 AEs was >5% for fatigue, thrombocytopenia, anemia and hand foot reaction and Incidence of Grade 3-4 AEs was >3% for fatigue, bilirubin elevation and anemia. Fluoropyrimidine-based chemotherapy was first provided to elderly individuals with advanced stomach cancer, and then metronomic capecitabine was given to the patients. According to the study, 2.2% and 13.3% of patients, respectively, had grade-II thrombocytopenia and neutropenia. The study found that non-hematologic toxicities of Grade II or III, such as stomatitis (13.4%), diarrhea (4.4%), and hand-foot syndrome (15.5%), were all reasonably common. Furthermore, no cases of treatment-related mortality, neutropenic fever, or Grade IV toxicity were reported ^39^. A study investigated the use of a metronomic capecitabine and cyclophosphamide treatment regimen for patients with inoperable or recurring pseudomyxoma peritonei. The findings suggest that this treatment strategy is well-tolerated with minimal side effects. Only a quarter (26%) of patients experienced moderate side effects (grade 3), and there were no instances of severe (grade 4) or life-threatening (grade 5) side effects ^40^. The pooled incidence of grade 3/4 AEs for metronomic chemotherapy regimens involving capecitabine, methotrexate, vinorelbine, bevacizumab etc. and its combinations of metastatic breast cancer was 29.5% (95% CI 21.1–39.5) ^38^.

Publication bias in the current study was measured using Egger’s test and funnel plots. Asymmetry in graphs was assessed using funnel plots. Only a small effect heterogeneity was observed, as indicated by p-values using Egger’s test. Regression of the standardised effect sizes on their precision is performed using Egger’s test; if publication bias is not present, the regression intercept should be zero. The weighted regression slope, and not the intercept, was predicted to be zero in the absence of publication bias. It is taken into account that this regression is similar to a weighted regression of the effect sizes on their standard errors, weighted by the inverse of their variances ^41^.

Sensitivity analyses focus on determining the degree of publication bias to measure the amount of bias that would be needed to reduce the observed point estimate to the null or to a selected non-null value, or its lower confidence interval (CI). This change in emphasis makes it possible to report relatively simple claims about susceptibility to publication bias^42^. Sensitivity analysis in this study indicated that the overall effect size remained statistically significant and directionally consistent with the original analysis.

There is a notable absence of universally accepted criteria and guidelines pertaining to the identification of suitable patient populations for metronomic chemotherapy, the selection of ideal dosage regimens, creation of personalized treatment schedules and the timing of use. Further clinical trials are required for evaluating the effectiveness of capecitabine.

More insightful approach can be made by comparing the effectiveness and safety of best supportive care and metronomic capecitabine. This would prove to be effective in giving future clinical discretion. Future studies should be carried out for the investigation of the combination therapies including conventional chemotherapy, radiotherapy and further other therapies related with exploration of biomarkers.

Although, through this meta-analysis we have summarized that metronomic capecitabine can have a significant effect in reducing hepatocellular carcinoma but there are some limitations of the study: firstly, the number of studies available for this analysis is quite less, hence, the study population is very low. Secondly, all the studies taken are non-RCT and are having less sufficient data. Thirdly, in some studies, data for PFS, OS, ORR, DCR and AEs are not available and cannot be extracted. Fourthly, we have taken the studies that were published in English language and the studies which were freely available to us. Nevertheless, to the best of our knowledge, this is first meta-analysis reporting effectiveness and safety of metronomic capecitabine in hepatocellular carcinoma.

## 5. Conclusions

Metronomic chemotherapy of capecitabine may be effectively used in patients with advanced hepatocellular carcinoma. Metronomic chemotherapy can be used alone and can be considered as an effective option as compared to combination therapies. More RCTs are required to be carried out so that more evidence could be obtained for the effectiveness of therapy of metronomic capecitabine on advanced hepatocellular cancer.

## Data Availability

All data produced in the present study are available upon reasonable request to the authors.

## Author contribution statement

All authors contributed equally to this manuscript.

*Nandini Gupta (NG):* Conceptualization, Methodology, Software, Data Extraction, Data curation, Writing-Original draft

*Neelkant Verma (NV):* Data Extraction, Data curation, Writing-Review & editing

*Bhoomika Patel (BP):* Conceptualization, Writing-Review & editing, Supervision, Final approval of manuscript.

## Ethics approval statement

This manuscript is a meta-analysis and not required an ethical approval.

## Patient consent statement

Not applicable

## Permission to reproduce material from other sources

Not applicable

## Clinical trial registration

Not applicable.

## Data availability statement

Data will be available upon reasonable request.

## Acknowledgements

Not applicable

## Abbreviations

CAPE: capecitabine
RCT: Randomized Control Trial
n-RCT: non-Randomized Control Trial
PFS: Progression Free Survival
OS: Overall Survival
CI: Confidence Interval
HCC: Hepatocellular carcinoma
SEER: Surveillance, Epidemiology, and End Results
MCT: Metronomic chemotherapy
DCR: Disease Control Rate
ORR: Overall Response Rate
ECOG score: Eastern Cooperative Oncology Group
m-RECIST: modified-Response Evaluation Criteria in Solid Tumors
RECIST: Response Evaluation Criteria in Solid Tumors
CR: Complete Response
PR: Partial response
SD: Stable Disease
MINORS: Methodological Index For Non-Randomized Studies.

## References

1. Peeters F, Dekervel J. Considerations for individualized first-line systemic treatment in advanced hepatocellular carcinoma. Curr Opin Pharmacol. 2023;70:102365. doi:10.1016/j.coph.2023.102365

2. Rumgay H, Arnold M, Ferlay J, et al. Global burden of primary liver cancer in 2020 and predictions to 2040. J Hepatol. 2022;77(6):1598–1606. doi:10.1016/j.jhep.2022.08.021

3. Liver Cancer Survival Rates | Cancer of the Liver Survival Rates | American Cancer Society. Accessed July 19, 2023. https://www.cancer.org/cancer/types/liver-cancer/detection-diagnosis-staging/survival-rates.html

4. Suresh D, Srinivas AN, Prashant A, Harikumar KB, Kumar DP. Therapeutic options in hepatocellular carcinoma: a comprehensive review. Clin Exp Med. Published online February 13, 2023. doi:10.1007/s10238-023-01014-3

5. Ikeda M, Morizane C, Ueno M, Okusaka T, Ishii H, Furuse J. Chemotherapy for hepatocellular carcinoma: current status and future perspectives. Jpn J Clin Oncol. 2018;48(2):103–114. doi:10.1093/jjco/hyx180

6. Sharma A, Houshyar R, Bhosale P, Choi JI, Gulati R, Lall C. Chemotherapy induced liver abnormalities: an imaging perspective. Clin Mol Hepatol. 2014;20(3):317. doi:10.3350/cmh.2014.20.3.317

7. Housman G, Byler S, Heerboth S, et al. Drug Resistance in Cancer: An Overview. Cancers (Basel). 2014;6(3):1769–1792. doi:10.3390/cancers6031769

8. Definition of metronomic chemotherapy - NCI Dictionary of Cancer Terms - NCI. Accessed April 12, 2023. https://www.cancer.gov/publications/dictionaries/cancer-terms/def/metronomic-chemotherapy

9. Maiti R. Metronomic chemotherapy. J Pharmacol Pharmacother. 2014;5(3):186-192. doi:10.4103/0976-500X.136098

10. Bahl A, Bakhshi S. Metronomic Chemotherapy in Progressive Pediatric Malignancies: Old Drugs in New Package. The Indian Journal of Pediatrics. 2012;79(12):1617–1622. doi:10.1007/s12098-012-0759-z

11. Cazzaniga ME, Cordani N, Capici S, Cogliati V, Riva F, Cerrito MG. Metronomic Chemotherapy. Cancers (Basel). 2021;13(9):2236. doi:10.3390/cancers13092236

12. Simsek C, Esin E, Yalcin S. Metronomic Chemotherapy: A Systematic Review of the Literature and Clinical Experience. J Oncol. 2019;2019:1–31. doi:10.1155/2019/5483791

13. Knikman JE, Rosing H, Guchelaar H, Cats A, Beijnen JH. A review of the bioanalytical methods for the quantitative determination of capecitabine and its metabolites in biological matrices. Biomedical Chromatography. 2020;34(1). doi:10.1002/bmc.4732

14. Xeloda | European Medicines Agency. Accessed July 25, 2023. https://www.ema.europa.eu/en/medicines/human/EPAR/xeloda

15. Liu J, He M, Wang Z, Li Q, Xu B. Current Research Status of Metronomic Chemotherapy in Combination Treatment of Breast Cancer. Oncol Res Treat. 2022;45(11):681–692. doi:10.1159/000526481

16. Li Q, He M, Liu J, Wang Z. Safety and efficacy study of oral metronomic capecitabine in combination with pyrotinib for HER2-positive metastatic breast cancer: A phase II trial. Journal of Clinical Oncology. 2023;41(16_suppl):e13038-e13038. doi:10.1200/JCO.2023.41.16_suppl.e13038

17. El-Sukhun S, Khalidi K. Durable effect of imatinib and metronomic chemotherapy with capecitabine in pancreatic carcinoma. Ecancermedicalscience. 2023;17. doi:10.3332/ecancer.2023.1535

18. Wang-Yuan Z, Jiang-Zheng Z, Lu Y Da, et al. Clinical efficacy of metronomic chemotherapy after cool-tip radiofrequency ablation in the treatment of hepatocellular carcinoma. International Journal of Hyperthermia. 2016;32(2):193–198. doi:10.3109/02656736.2015.1099168

19. Granito A, Marinelli S, Terzi E, et al. Metronomic capecitabine as second-line treatment in hepatocellular carcinoma after sorafenib failure. Digestive and Liver Disease. 2015;47(6):518–522. doi:10.1016/j.dld.2015.03.010

20. De Lorenzo S, Tovoli F, Barbera MA, et al. Metronomic capecitabine vs. best supportive care in Child-Pugh B hepatocellular carcinoma: a proof of concept. Scientific Reports (Nature Publisher Group). 2018;8:1–7. doi:10.1038/s41598-018-28337-6

21. Casadei Gardini A, Foca F, Scartozzi M, et al. Metronomic capecitabine versus best supportive care as second-line treatment in hepatocellular carcinoma: a retrospective study. Scientific Reports (Nature Publisher Group). 2017;7:42499. doi:10.1038/srep42499

22. Brandi G, Rosa F, Agostini V, et al. Metronomic Capecitabine in Advanced Hepatocellular Carcinoma Patients: A Phase II Study. Oncologist. 2013;18(12):1256–1257. doi:10.1634/theoncologist.2013-0093

23. Pelizzaro F, Gambato M, Gringeri E, et al. Management of Hepatocellular Carcinoma Recurrence after Liver Transplantation. Cancers (Basel). 2021;13(19):4882. doi:10.3390/cancers13194882

24. Trevisani F, Brandi G, Garuti F, et al. Metronomic capecitabine as second-line treatment for hepatocellular carcinoma after sorafenib discontinuation. J Cancer Res Clin Oncol. 2018;144(2):403–414. doi:10.1007/s00432-017-2556-6

25. Ravaioli M, Cucchetti A, Pinna AD, et al. The role of metronomic capecitabine for treatment of recurrent hepatocellular carcinoma after liver transplantation. Scientific Reports (Nature Publisher Group). 2017;7:1–9. doi:10.1038/s41598-017-11810-z

26. Peristeri D V, Tepelenis K, Karampa A, et al. Metronomic chemotherapy with cyclophosphamide for the treatment of advanced hepatocellular cancer: A case report. Ann Med Surg (Lond). 2021;72:103043. doi:10.1016/j.amsu.2021.103043

27. Torimura T, Iwamoto H, Nakamura T, et al. Metronomic chemotherapy: possible clinical application in advanced hepatocellular carcinoma. Transl Oncol. 2013;6(5):511–519. doi:10.1593/tlo.13481

28. Yang H, Woo HY, Lee SK, et al. A comparative study of sorafenib and metronomic chemotherapy for Barcelona Clinic Liver Cancer-stage C hepatocellular carcinoma with poor liver function. Clin Mol Hepatol. 2017;23(2):128–137. doi:10.3350/cmh.2016.0071

29. Banchi M, Lanzolla T, Di Napoli A, Bandini A, Bocci G, Cox MC. Complete remission of a diffuse large B-cell lymphoma in a young patient, with severe tuberous sclerosis, treated with metronomic chemotherapy and ibrutinib: a case report. Chemotherapy. Published online August 7, 2023. doi:10.1159/000533236

30. Mireștean CC, Iancu RI, Iancu DPT. Capecitabine-A “Permanent Mission” in Head and Neck Cancers “War Council”? J Clin Med. 2022;11(19). doi:10.3390/jcm11195582

31. Schellens JHM. Capecitabine. Oncologist. 2007;12(2):152-155. doi:10.1634/theoncologist.12-2-152

32. Zafar SY, Hirsch. Capecitabine in the management of colorectal cancer. Cancer Manag Res. Published online March 2011:79. doi:10.2147/CMR.S11250

33. Geng R, Wang G, Qiu L, et al. Metronomic capecitabine as maintenance treatment after first line induction with XELOX for metastatic colorectal cancer patients. Medicine. 2020;99(51):e23719. doi:10.1097/MD.0000000000023719

34. Berruti A, Fazio N, Ferrero A, et al. Bevacizumab plus octreotide and metronomic capecitabine in patients with metastatic well-to-moderately differentiated neuroendocrine tumors: the xelbevoct study. BMC Cancer. 2014;14(1):184. doi:10.1186/1471-2407-14-184

35. Ibrahim T, Bongiovanni A, Riva N, et al. Metronomic capecitabine in gastroenteropancreatic neuroendrocrine tumors: a suitable regimen and review of the literature. Onco Targets Ther. Published online October 2014:1919. doi:10.2147/OTT.S68573

36. Fedele P, Marino A, Orlando L, et al. Efficacy and safety of low-dose metronomic chemotherapy with capecitabine in heavily pretreated patients with metastatic breast cancer. Eur J Cancer. 2012;48(1):24–29. doi:10.1016/j.ejca.2011.06.040

37. Yuan Z, Hua X, Li WZ, et al. Predict the benefit of metronomic capecitabine maintenance in early-stage triple-negative breast cancer: Results from the SYSUCC- 001 study. Journal of Clinical Oncology. 2021;39(15_suppl):521-521. doi:10.1200/JCO.2021.39.15_suppl.521

38. Liu Y, Gu F, Liang J, et al. The efficacy and toxicity profile of metronomic chemotherapy for metastatic breast cancer: A meta-analysis. PLoS One. 2017;12(3):e0173693. doi:10.1371/journal.pone.0173693

39. He S, Shen J, Hong L, Niu L, Niu D. Capecitabine “metronomic” chemotherapy for palliative treatment of elderly patients with advanced gastric cancer after fluoropyrimidine-based chemotherapy. Medical Oncology. 2012;29(1):100–106. doi:10.1007/s12032-010-9791-x

40. Raimondi A, Corallo S, Niger M, et al. Metronomic Capecitabine With Cyclophosphamide Regimen in Unresectable or Relapsed Pseudomyxoma Peritonei. Clin Colorectal Cancer. 2019;18(2):e179–e190. doi:10.1016/j.clcc.2019.03.002

41. Lin L, Chu H. Quantifying Publication Bias in Meta-Analysis. Biometrics. 2018;74(3):785–794. doi:10.1111/biom.12817

42. Mathur MB, VanderWeele TJ. Sensitivity Analysis for Publication Bias in Meta-Analyses. J R Stat Soc Ser C Appl Stat. 2020;69(5):1091–1119. doi:10.1111/rssc.12440

